# Effect of comprehensive medication management on mortality in critically ill patients

**DOI:** 10.1101/2024.10.23.24316000

**Authors:** Andrea Sikora, Wanyi Min, Mengxuan Hu, John W. Devlin, David J. Murphy, Brian Murray, Bokai Zhao, Ye Shen, Xianyan Chen, Susan E. Smith, Sandra Rowe, Tianming Liu, Sheng Li

**Author notes:** 12850 E Montview Blvd, Aurora, CO 80045 | Mail Stop C238. On behalf of the MRC-ICU Investigator Team. **Funding:** Funding through Agency of Healthcare Research and Quality for Drs. Devlin, Murphy, Sikora, Smith, Shen, Li, Liu, and Kamaleswaran was provided through R21HS028485 and R01HS029009.

## Abstract

**Background:** Medication management in the intensive care unit (ICU) is causally linked to both treatment success and potential adverse drug events (ADEs), often associated with deleterious consequences. Patients with higher severity of illness tend to require more management. The purpose of this evaluation was to explore the effect of comprehensive medication management (CMM) on mortality in critically ill patients.

**Methods:** In this retrospective cohort study of adult ICU patients, CMM was measured by critical care pharmacist (CCP) medication interventions. Propensity score matching was performed to generate a balanced 1:1 matched cohort, and logistic regression was applied for estimating propensity scores. The primary outcome was the odds of hospital mortality. Hospital and ICU length of stay were also assessed.

**Results:** In a cohort of 10,441 ICU patients, the unadjusted mortality rate was 11% with a mean APACHE II score of 9.54 ± 4.18 and Medication Regimen Complexity-Intensive Care Unit (MRC-ICU) score of 5.78 ± 4.09. Compared with CCP interventions less than 3, more CCP interventions was associated with a significantly reduced risk of mortality (estimate -0.04, 95% confidence interval -0.06 - -0.03, p < 0.01) and shorter length of ICU stay (estimate -2.77, 95% CI -2.98 - - 2.56, p < 0.01).

**Conclusions:** The degree by which CCPs deliver CMM in the ICU is directly associated with reduced hospital mortality independent of patient characteristics and medication regimen complexity.

## Introduction

The participation of critical care pharmacists (CCPs) on multiprofessional intensive care unit (ICU) rounds are associated with a nearly 70% reduction in adverse drug events (ADEs) and a 22% reduction in mortality.^1-3^ These benefits are thought to be conferred by the cognitively-intense direct patient care CCPs provide during the comprehensive medication management (CMM) process.^4^ CMM has been defined as “a patient-centered approach to optimizing medication use and improving patient health outcomes that is delivered by a clinical pharmacist working in collaboration with the patient and other healthcare providers.”^5^

The most common strategies to measure CMM services in the ICU include the number of patients being provided with CMM and the number of CCP interventions made.^6^ Another approach is to quantify the complexity of the medication regimen of each individual patient, as a proxy towards the cognitive load required by the CCP to adequately assess, monitor, and effect change upon those medication regimens through interventions. The Medication Regimen Complexity-Intensive Care Unit (MRC-ICU) score has been associated with patient-centered outcomes (mortality, length of stay), ICU complications (drug-drug interactions, prolonged duration of mechanical ventilation, fluid overload), and pharmacist workload (interventions, orders verified).^7-17^ Current research suggests medication regimen complexity parallels illness severity and that an increasing MRC-ICU is associated with higher rates of mortality and other worsening outcomes as well as a higher CCP workload. These assumptions may give the potentially false impression that greater CMM is associated with worse patient outcome(s). However, in all likelihood, a causal pathway may exist connecting ICU patient characteristics (including severity of illness), medication regimen complexity, the scope of CMM delivered, and patient outcome(s). A causal framework for this proposed pathway is summarized in **Figure 1**.

**Figure 1.**
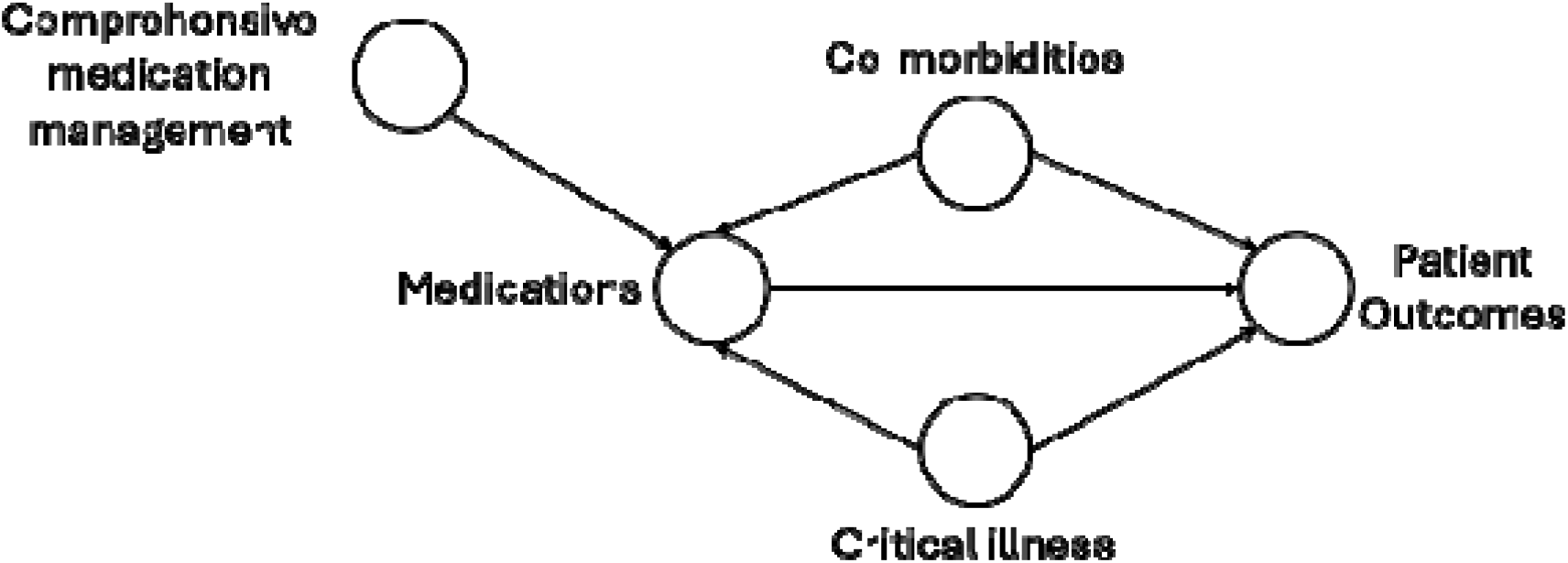
Directed acyclic graph for the causal pathway relating comprehensive medication management to medications that patients receive and patient outcomes.

The purpose of this study is to formally evaluate this potential causal pathway in an effort to better evaluate the effect of CCP-delivered CMM on patient-centered outcomes, including hospital mortality. We hypothesized that patients who received more CMM services, as measured by CCP interventions, would have lower mortality compared to patients receiving fewer CMM services but with similar baseline characteristics and medication regimen intensities.

## Methods

### Study Design

This evaluation was a retrospective, observational cohort study that employed propensity score matching to evaluate the potential causal role of CCP medication interventions during the CMM process on hospital mortality (primary outcome), ICU length of stay, and hospital length of stay. The independent variable of interest was the total medication intervention count as logged in Epic^®^ as i-Vents by a CCP during the patient’s ICU hospitalization.

Interventions were totaled for both days spent in the ICU and non-ICU hospital settings.

This study was reviewed by the University of Georgia (UGA) Institutional Review Board (IRB) and determined to be exempt from IRB oversight (Project00001541). All methods were performed in accordance with the ethical standards of the UGA IRB and the Helsinki Declaration of 1975.

### Study Population

Data were obtained by the Oregon Clinical and Translational Research Institute, which manages Epic^®^ electronic health record (EHR) data from Oregon Health and Science University (OHSU) Hospital. Data extraction included a total of 13,500 patients aged 18 years or older who were admitted to an ICU between June 1, 2020 and June 7, 2023. Data from only the index ICU admission for each patient were included. Patients were excluded if the ICU stay was less than 24 hours or if the patient was placed on comfort care based on Epic^©^ order-sets validated by a local clinician within the first 24 hours of the ICU stay.

### Variables

The EHR was queried for patient demographic information, medication information, and patient outcomes. Patient characteristics included age, sex, race, ICU type, admission diagnosis, APACHE II score 24 hours after ICU admission, SOFA score 24 hours after ICU admission, and the MRC-ICU score 24 hours, 48 hours, and 72 hours after ICU admission. The MRC-ICU consists of 35 discrete medication categories with each category assigned a weighted value that is then summed to create a score for a patient’s regimen at the given time point. For instance, if a patient was prescribed vancomycin, norepinephrine, and insulin, they would be given 3 points for vancomycin, 1 point for norepinephrine, and 1 point for insulin for a total score of 5. The MRC-ICU score has been previously built into the EHR at OHSU.^16,17^ Patient outcomes included hospital mortality, ICU length of stay, and hospital length of stay.

### Pre-processing

**Supplemental Figure 1** provides a CONSORT diagram for the data included in this analysis. One-hot encodings for the categorical variables such as ICU type and ICU admission diagnosis were created, representing these variables as numerical values for further analysis.

### Propensity score matching

To reduce the treatment assignment bias and mimic randomization, propensity score matching was applied. This method is intended to reduce the effects of confounding variables in observational studies by creating two comparable groups of subjects between the treated (e.g., high intervention exposure) and control (low intervention exposure) groups.^18^

The treatment variable was the total medication intervention count during the patient’s ICU stay and during the entire hospital stay. These were explored at a threshold of 3 interventions for the ICU stay and 3 and 5 interventions for hospital stay (see **Supplemental Figure 2**). A patient exceeding these thresholds was assigned to the treatment group and under these thresholds was assigned to the control group. A secondary analysis that evaluated total hospital interventions for an ICU patient during their hospitalization used the same threshold (3 and 5 were again used) for assignment to the treatment and control groups.

Logistic regression was used for estimating propensity scores, where the binary target of the probability was whether the case received treatment (i.e., exceeded high CCP intervention threshold) or not (i.e., below high CCP intervention threshold), and the predictors were the covariates (i.e., admission variables, 24-hour variables) on which the cases were to be matched. Each patient in the treated group was matched 1:1 with a patient in the control group. The matching process ensured that each case from the non-treatment cohort had its closest counterpart in the treatment group based on the calculated propensity score using the K-Nearest Neighbors (K-NN) algorithm.

### Average treatment effect

After propensity score matching, the average treatment effect (ATE) of the two treatments on each outcome were calculated. The ATE, often used to measure the average effect of an intervention or treatment across a population, quantifies the difference in the expected outcomes if all individuals in the study population were treated versus if all individuals were not treated.^19^ A positive ATE value indicates that the treatment has a beneficial effect on the outcome, a negative ATE value indicates that the treatment has a detrimental effect on the outcome, and zero ATE indicates that the treatment has no effect on the outcome. The changes in these effects before and after matching were visualized to help confirm whether the matching process effectively balanced the differences between the treatment and control groups, thereby making subsequent analyses more reliable.

### Analysis

Descriptive statistics were performed for relevant variables. Continuous variables were summarized by the mean and standard deviation, and categorical variables reported count and proportion of the total population. The t-test was adopted, and a two-sided p-value less than 0.05 was used to determine statistical significance for all outcomes. All analyses were performed using *R* (version 4.1.2).

## Results

The initial cohort was 13,500 ICU patients with 3,059 excluded due to incomplete relevant data for a total of 10,441 patients (see **Supplemental Table 1**). The average age was 59.3 ± 17.6 years with a 24-hour APACHE II score of 9.54 ± 4.18 and MRC-ICU score of 5.78 ± 4.09. The rate of mortality was 11% with an average length of ICU stay of 5.19 ± 6.43. Patient information is provided in **Table 1**.

**Table 1.**
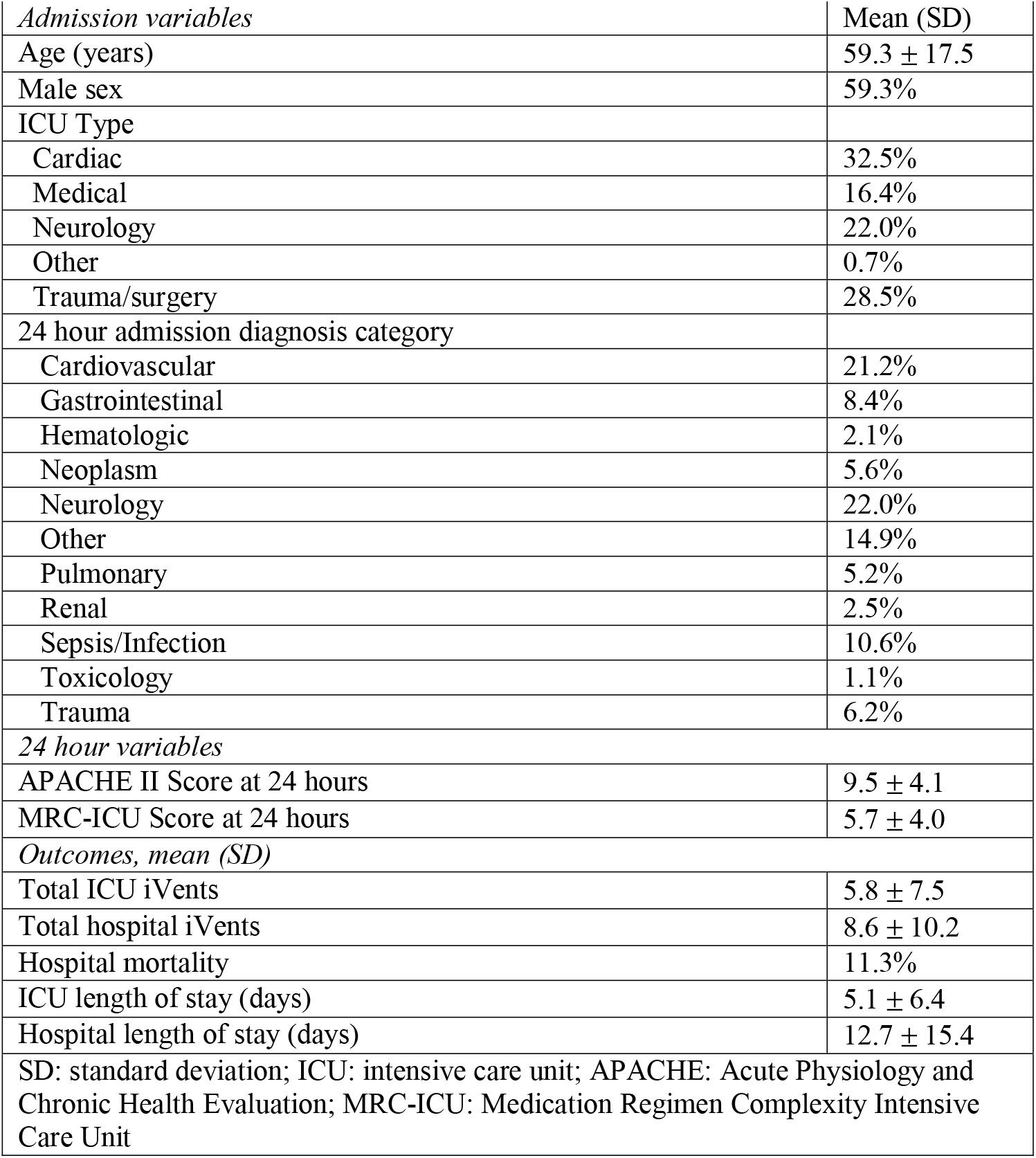
Patient information and outcomes.

For ICU interventions at a threshold of 3, there were 4,029 pairs of rows matched. The propensity score matching process improved the balance between the treatment and control groups, as evidenced by the substantial reduction (below 0.1) in effect sizes for nearly all covariates (**Figure 2**). Initially, the ATE of mortality, ICU length of stay, and hospital length of stay were +0.08, +3.82, and -0.44, indicating that mortality and ICU length of stay in the treatment groups were higher than those in the control groups. However, after matching, the ATE of mortality and ICU length of stay fell below zero (−0.04 and -2.77), while hospital length of stay increased above zero (0.49). These are summarized in **Figure 3**.

**Figure 2.**
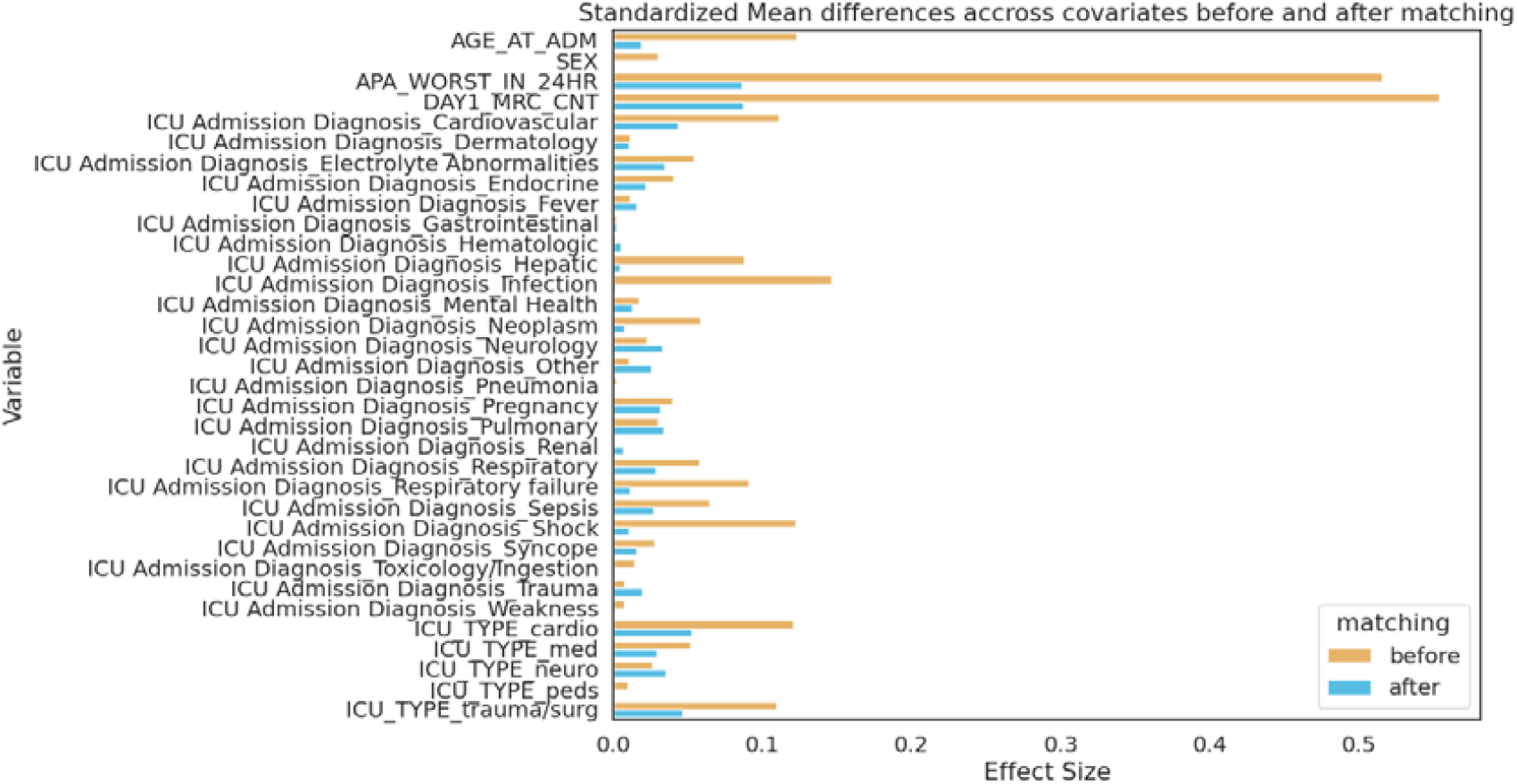
Standardized mean differences across variables before and after matching for ICU intervention with threshold 3.

**Figure 3a.**
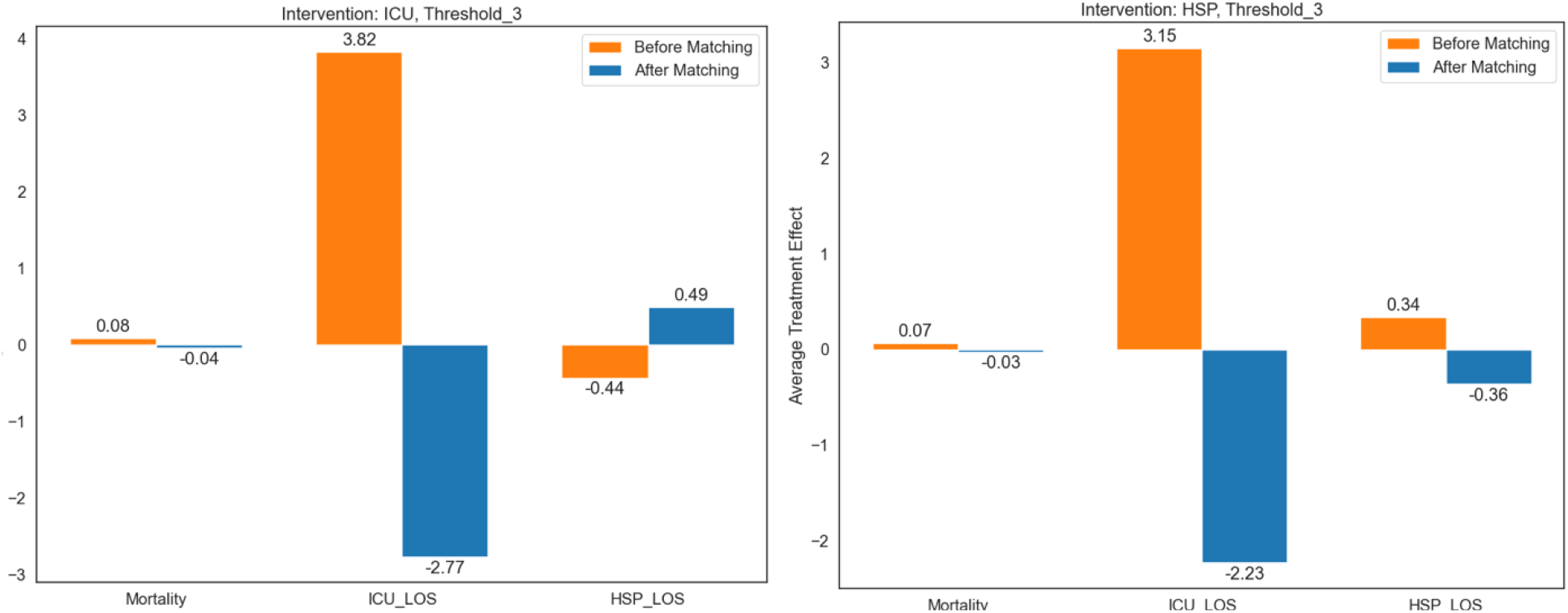
ATE values before (left) and after (right) matching for ICU intervention with threshold 3. **Figure 3b**. ATE values before (left) and after (right) matching for hospital intervention with threshold 3.

For hospital interventions at a threshold of 3, there were 2,103 patients matched (**Supplemental Figure 3**). Before matching, the ATE of mortality, ICU length of stay, and hospital length of stay were 0.07, 3.15 and 0.34. After propensity score matching, the ATE of mortality was -0.03, ICU length of stay was -2.23, and hospital length of stay was -0.36 (**Figure 4**). Thresholds of 5 were also explored, but the persistence of large effect sizes (exceeding 0.2) for some covariates post-matching indicates that residual confounding remains (**Supplemental Figures 4-5**).

**Table 2** summarizes the propensity-matched analysis for CCP interventions. After conducting propensity score matching at a ICU intervention threshold of 3, patients receiving more CCP interventions in the ICU exhibited reduced mortality compared to non-receivers (ATE = -0.04; CI, -0.06 to -0.03; P < 0.05). Additionally, those receiving more ICU interventions had a shorter ICU length of stay (ATE = -2.77; CI, -2.98 to -2.56; P < 0.05) while hospital length of stay showed no significant difference (ATE = 0.49; CI, -0.19 to 1.17; P = 0.16). Similar trends where the ATE was reversed were observed for hospital interventions at thresholds of 3 and 5.

**Table 2.** Results of propensity-matched analysis.

| **Table 2. Results of propensity-matched analysis** |  |  |  |
| --- | --- | --- | --- |
| *Pharmacist interventions >3 during the ICU stay (n = 4,029)* |  |  |  |
|  | ATE | p-value | 95% Confidence Interval |
| Mortality | -0.04 | <0.01 | -0.06 to -0.03 |
| ICU length of stay | -2.77 | <0.01 | -2.98 to -2.56 |
| Hospital length of stay | 0.49 | 0.16 | -0.19 to 1.17 |
| *Pharmacist interventions during the hospital stay >3 (n = 2,103)* |  |  |  |
| Mortality | -0.03 | <0.01 | -0.04 to -0.01 |
| ICU length of stay | -2.23 | <0.01 | -2.51 to -1.95 |
| Hospital length of stay | -0.36 | 0.44 | -1.28 to 0.55 |
| *Pharmacist interventions during the hospital stay >5 (n = 4,260)* |  |  |  |
| Mortality | -0.04 | <0.01 | -0.05 to -0.03 |
| ICU length of stay | -2.99 | <0.01 | -3.19 to -2.79 |
| Hospital length of stay | -0.08 | 0.81 | -0.75 to 0.58 |

## Discussion

After applying rigorous causal inference methods to critical care pharmacist delivery of comprehensive medication management, we observed patients receiving more pharmacist medication interventions had lower rates of mortality and reduced length of ICU stay after controlling for multiple patient factors, including severity of illness, and the intensity of the medication regimen on the first three ICU days. The results represent a reversal of the directionality of the relationship between CCP interventions and patient outcomes observed in prior analyses (add refs) where an increased number of CCP interventions appeared to be associated with increased rates of mortality and longer ICU length of stay.

While it can be postulated that patients with higher severity of illness require more intervention (both in the form of medications as well as surgeries, supportive care devices, etc.), these patients are also at a higher absolute risk of dying regardless of those interventions. Medications, which are independent risk factors for adverse outcomes, also constitute a significant realm of potential interventions in the management of critical illness and represent potentially life-saving treatments.^4^ This can paradoxically create an observed association in which it appears that higher medication regimen complexity and potentially higher amounts of medication interventions as performed by CCPs confer a higher risk of mortality.

The directed acyclic graph presented in **Figure 1** is based on a number of observational studies and previous conceptual frameworks.^4,20,21^ In particular, the value of CCPs and common ICU medication interventions on patient outcomes has been well documented. In 1999, the landmark trial by Leape et al. observed that pharmacists participating on multidisciplinary rounds were associated with a nearly 70% reduction in ADEs.^2^ In 2019, Lee et al. conducted a systematic review and meta-analysis and observed that pharmacists participating on multidisciplinary rounds were associated with substantial reductions in ADEs as well as reduced length of ICU stay (by approximately 1.2 days) and reduced odds of mortality (by approximately 20%).^3^ In 2018, Leguelinel-Blache et al. observed that a quality improvement oriented bundle that targeted a number of common interventions including antibiotic stewardship and sedation management was associated with reductions in hospital and ICU length of stay, duration of mechanical ventilation, as well as costs.^22^

Previous explorations of medication regimen complexity in the ICU have important causal implications. In a 28-center prospective observational study of 3,908 patients, increasing MRC-ICU was associated with both more pharmacist interventions and higher intervention intensity in addition to higher rates of mortality and longer lengths of stay.^23^ Interestingly, this study also found that in situations of higher workload (as measured by more ICU patients per pharmacist), there were fewer interventions and lower intervention intensity as well as longer length of stay. Although this study was not designed to draw causal conclusions, it suggested a possibility that as CCP workload increases, they are less able to do their roles effectively, which could potentially result in worse patient outcomes. Although these findings had face validity due to observations that CCPs on rounds are beneficial to outcomes, previous evaluations had found that higher MRC-ICU scores were linked with both higher CCP interventions as well as worse outcomes. Indeed, in most of the original studies of the MRC-ICU, a higher score was related to higher mortality and longer lengths of stay,^7,8,12,24,25^ In the first analysis to account for severity of illness, the relationship flipped directions, with higher MRC-ICU being related to lower rates of mortality. That analysis opened up the question of if there is a “Goldilocks effect” to medication regimen complexity, with sick patients needing “appropriately” complex regimens that balance the needs of that disease against the risks of the medication treatment.^26^

This study fills an important conceptual gap with the finding that in patients matched on MRC-ICU and severity of illness (i.e., APACHE II), more CCP interventions conferred improved outcomes. This finding has important clinical implications by showing that CMM services are an important way to improve care quality and safety, which is salient given that 30% of ICUs lack CCP/CMM services and many patients do not receive CMM on holidays or weekends.^27^ Moreover, high workload may reduce CMM quality, which now has important links to patient outcomes.

Limitations of this study include its single center, observational design. The evaluation including hospital interventions likely reflects the continuum of care provided by pharmacists (both in the ICU and on ward services), beyond just CCPs. Moreover, although interventions capture elements of the CCP’s direct patient care activities, it has been acknowledged that there are limitations to this singular measure, and future evaluations would ideally capture a more nuanced evaluation of CCP cognitive services as encompassed by CMM.^4,16,28^ However, it is not a common practice for CCPs to regularly document interventions, such that this dataset represents a highly unique glimpse into CCM as provided by CCPs. Also, in the future, large language models with advanced reasoning capabilities, such as the OpenAI o1 model, could be potentially employed to cross-verify the causal pathway and the results described in this paper.

## Conclusion

The use of propensity score matching in the context of a causal framework demonstrates the intensity of comprehensive medication management services delivered by critical care pharmacists is associated with improved patient-centered outcomes, including mortality.

## Data Availability

All data produced in the present study are available upon reasonable request to the authors

## Acknowledgements

Amoreena Most, PharmD, BCCCP; Aaron Chase, PharmD, BCCCP; OHSU Oregon Clinical and Translational Research Institute

## Supplemental Digital Content

**Supplemental Figure 1.**
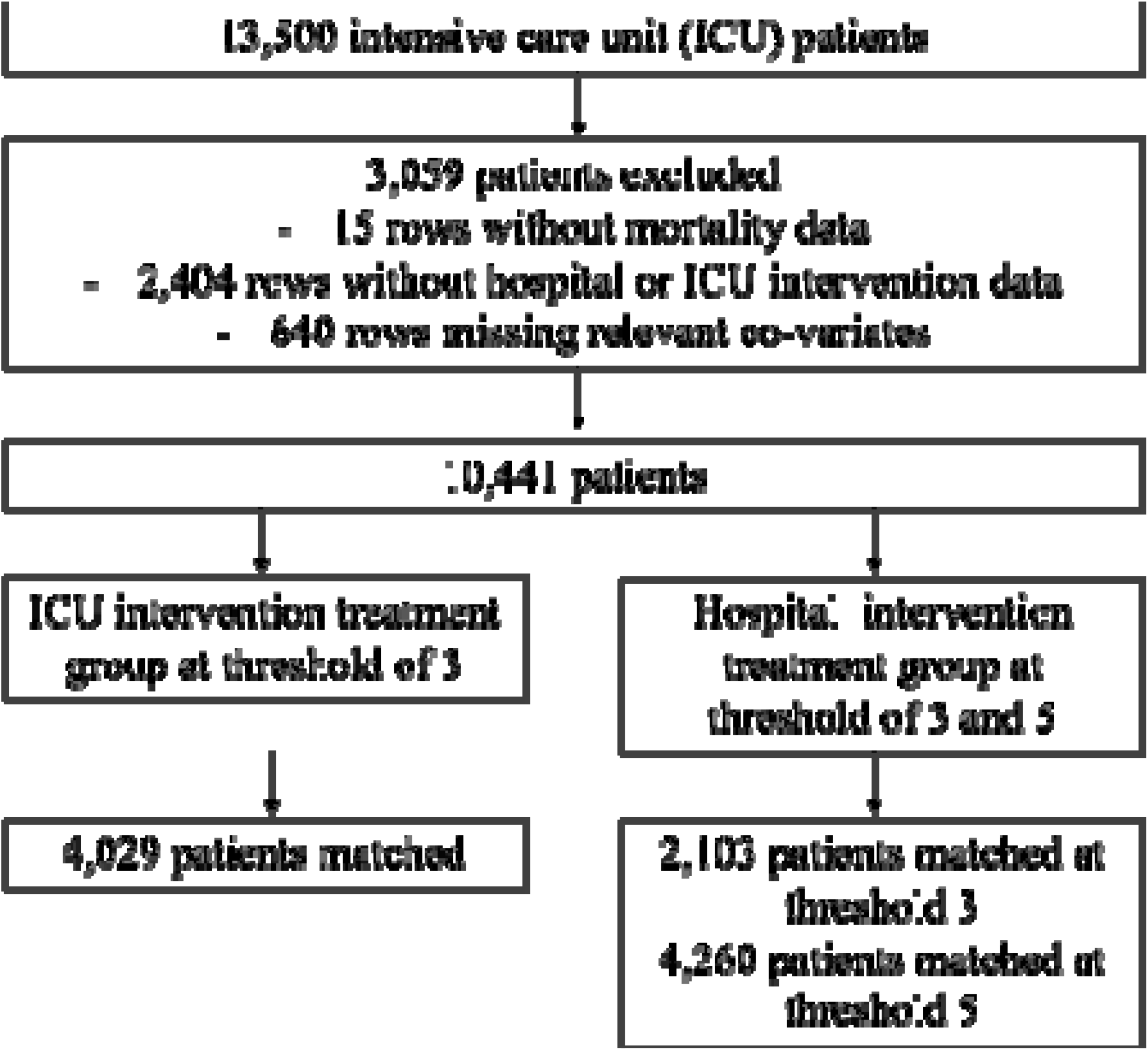
CONSORT Diagram.

**Supplemental Figure 2.**
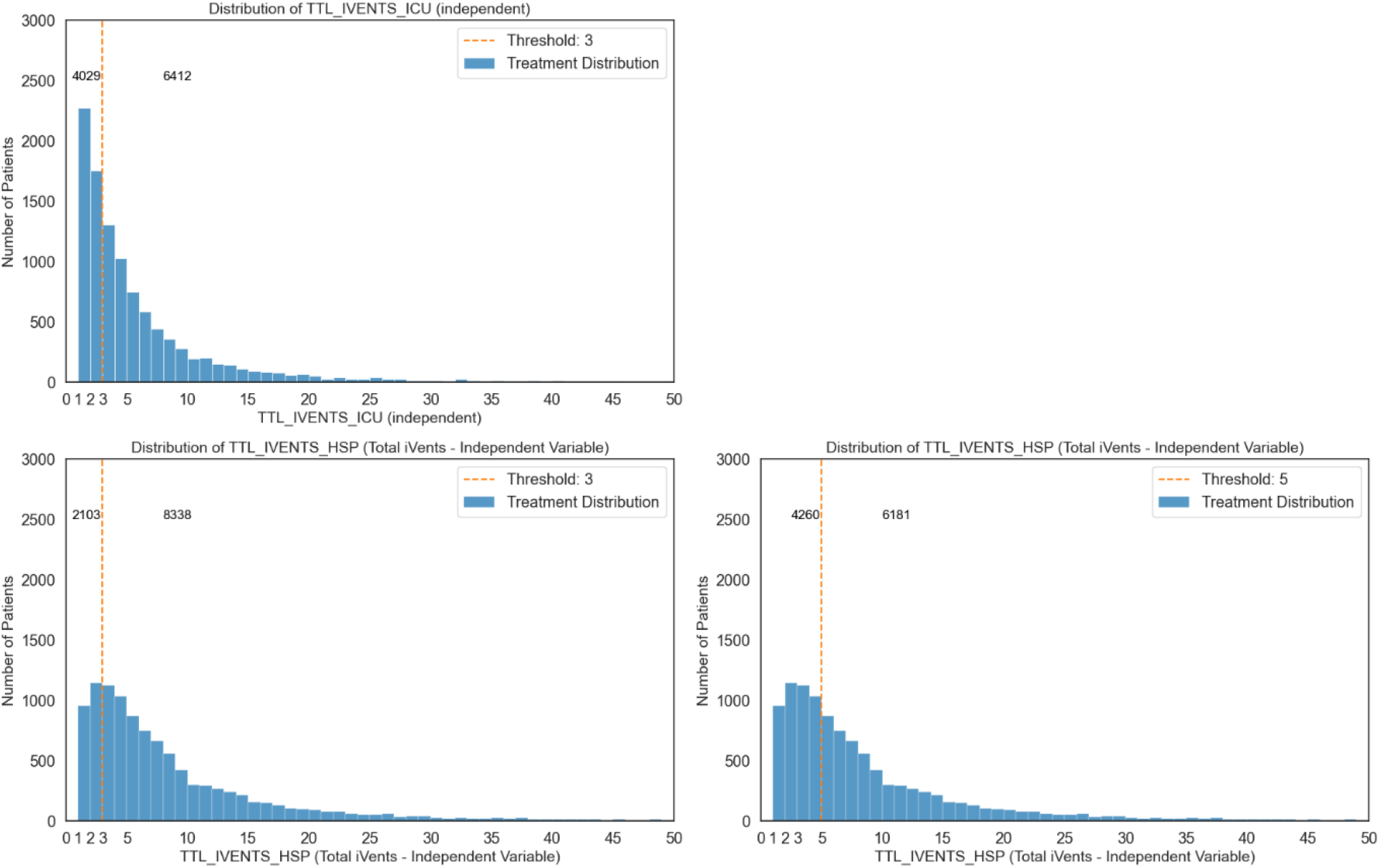
Histogram of Intervention Thresholds. The top panel indicates ICU interventions, and the bottom two panels indicate total hospital interventions. The histograms show the distribution of intervention counts for all patients, with the dotted line indicating the treatment threshold, where above this line was considered the active treatment group and below the line considered the control group for causal analysis.

**Supplemental Figure 3.**
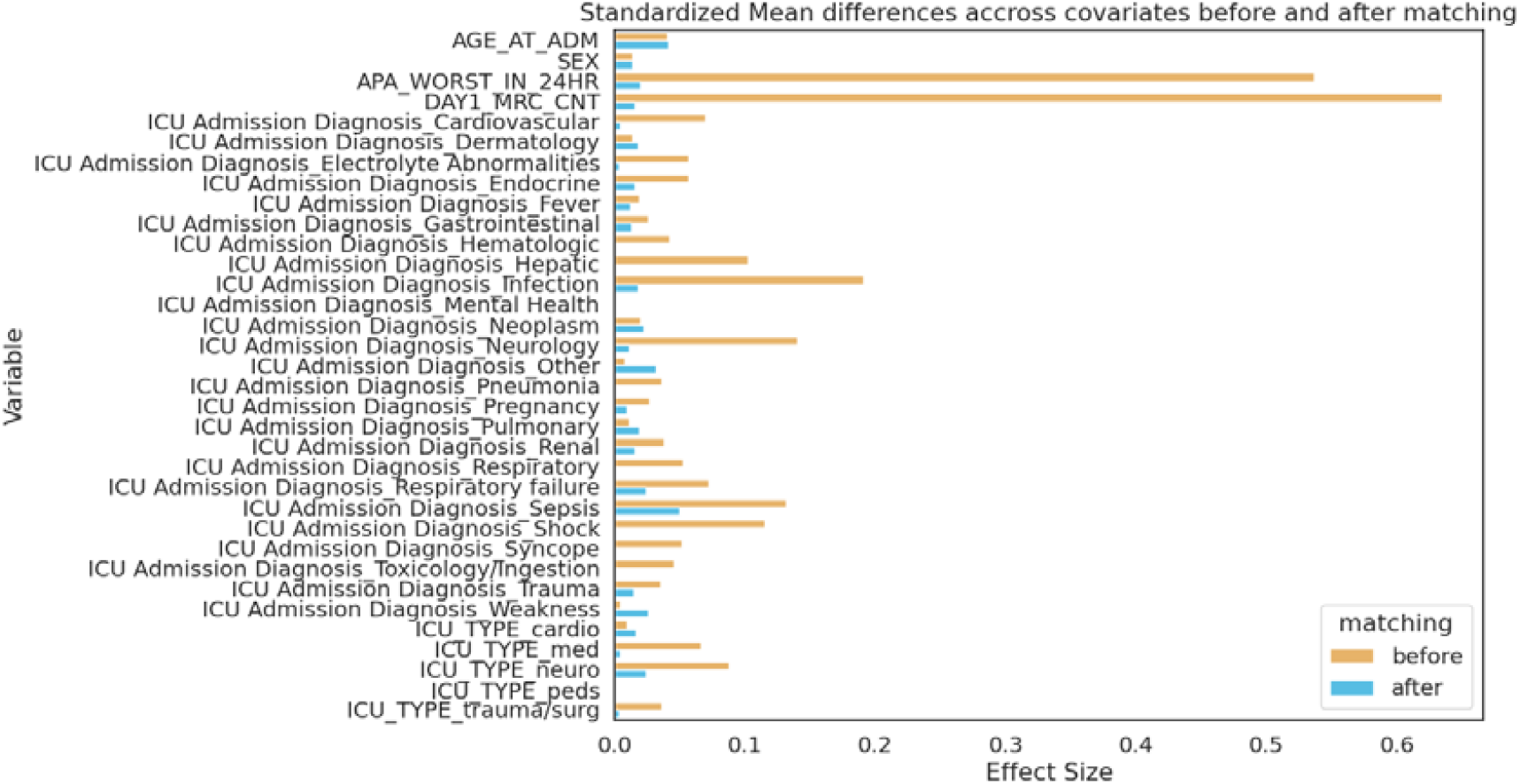
Standardized mean differences across variables before and after matching for hospital intervention with threshold 3.

**Supplemental Figure 4.**
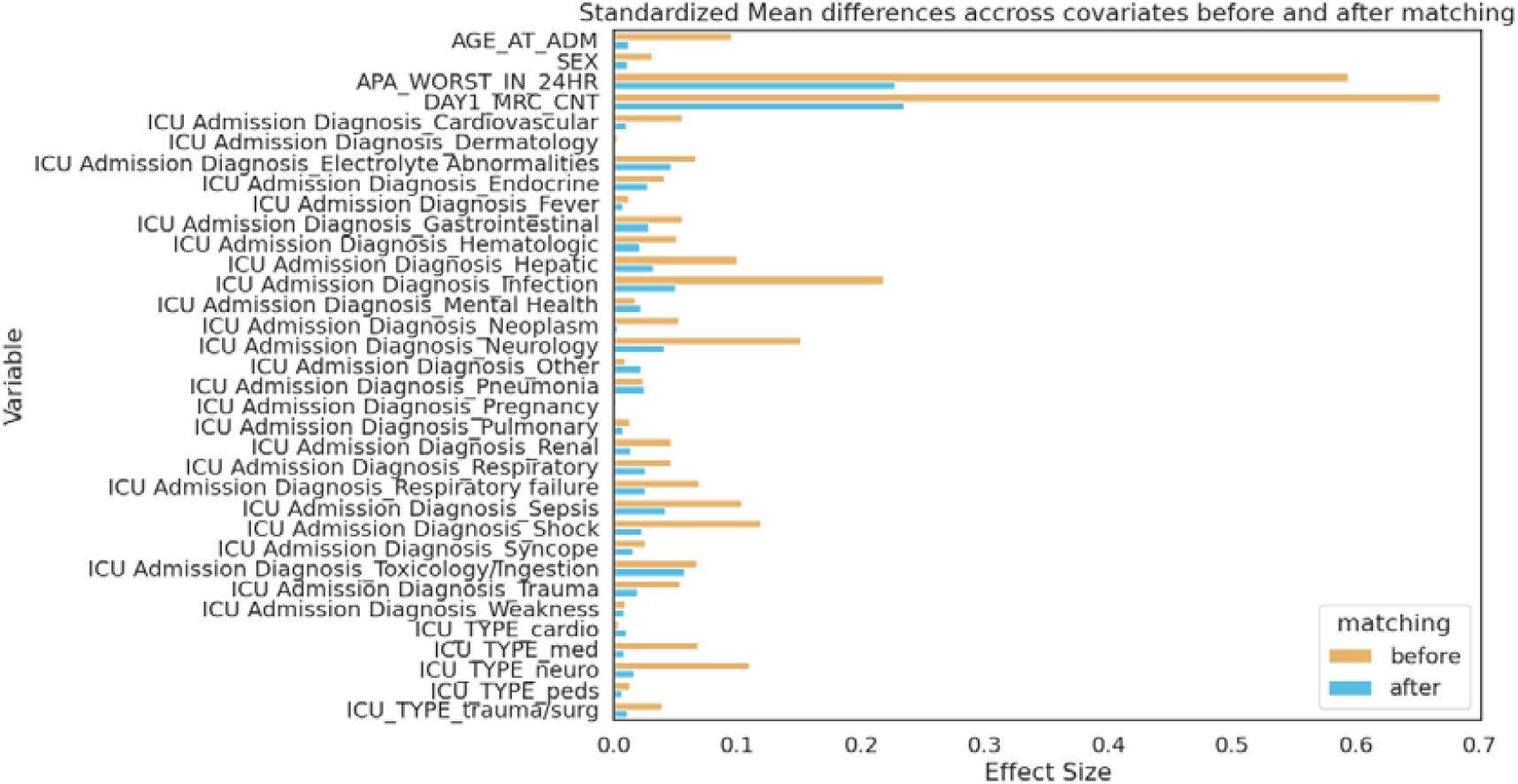
Standardized mean differences across variables before and after matching for hospital intervention with threshold 5.

**Supplemental Figure 5.**
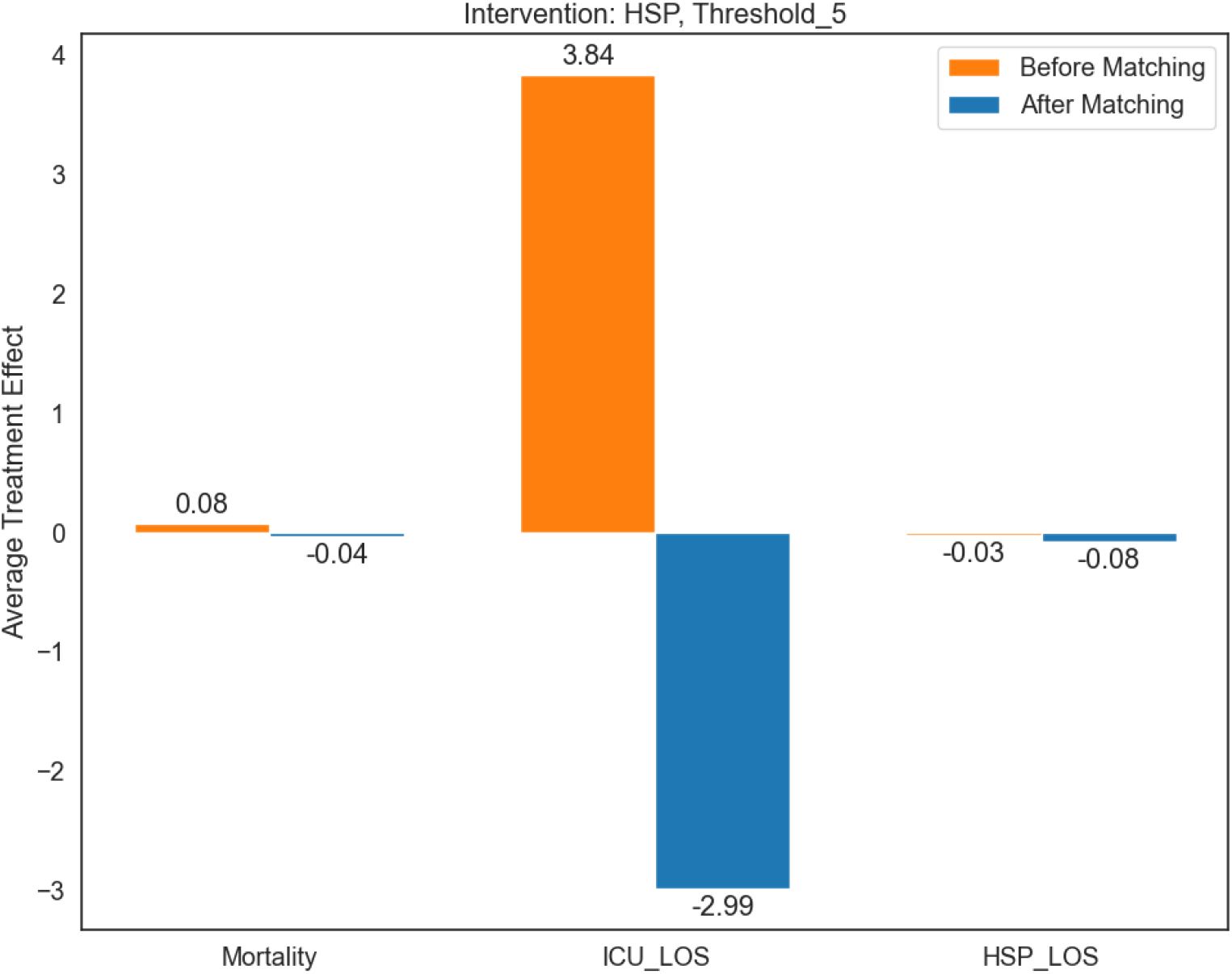
ATE values before (left) and after (right) matching for hospital intervention with threshold 5. There were 4,260 pairs of matching patients. The persistence of large effect sizes (exceeding 0.2) for some covariates post-matching indicates that residual confounding remains that may attribute to observed outcomes.

